# How often do cancer researchers make their data and code available and what factors are associated with sharing?

**DOI:** 10.1101/2022.03.10.22272231

**Authors:** Daniel G. Hamilton, Matthew J. Page, Sue Finch, Sarah Everitt, Fiona Fidler

## Abstract

**Background:** Various stakeholders are calling for increased availability of data and code from cancer research. However, it is unclear how commonly these products are shared, and what factors are associated with sharing. Our objective was to evaluate how frequently oncology researchers make data and code available, and explore factors associated with sharing.

**Methods:** A cross-sectional analysis of a random sample of 306 articles indexed in PubMed in 2019 presenting original cancer research was performed. Outcomes of interest included the prevalence of affirmative sharing declarations and the rate with which declarations connected to useable data. We also investigated associations between sharing rates and several journal characteristics (e.g., sharing policies, publication models), study characteristics (e.g., cancer rarity, study design), open science practices (e.g., pre-registration, pre-printing) and citation rates between 2020-2021.

**Results:** One in five studies declared data were publicly available (95% CI: 15-24%). However, when actual data availability was investigated this percentage dropped to 16% (95% CI: 12-20%), and then to less than 1% (95% CI: 0-2%) when data were checked for compliance with key FAIR principles. While only 4% of articles that used inferential statistics reported code to be available (10/274, 95% CI: 2-6%), the odds of reporting code to be available were 5.6 times higher for researchers who shared data. Compliance with mandatory data and code sharing policies was observed in 48% and 0% of articles, respectively. However, 88% of articles included data availability statements when required. Policies that encouraged data sharing did not appear to be any more effective than not having a policy at all. The only factors associated with higher rates of data sharing were studying rare cancers and using publicly available data to complement original research.

**Conclusions:** Data and code sharing in oncology occurs infrequently, and at a lower frequency than would be expected due to non-compliance with journal policies. There is also a large gap between those declaring data to be available, and those archiving data in a way that facilitates its reuse. We encourage journals to actively check compliance with sharing policies, and researchers consult community accepted guidelines when archiving the products of their research.

---

The notion that scientific findings should be independently verifiable is a key tenet of science, with this principle famously being enshrined in the motto of the Royal Society of London in 1660. However, the extent to which researchers adhere to this value in practice varies across fields.

In the context of contemporary medical research, the inability to establish the veracity of important findings can cast doubts on the validity of research, and sometimes lead to retraction.^1-4^ Perhaps the most recent and well-known example being the retraction of two influential papers that investigated the effectiveness of the antimalarial drugs chloroquine and hydroxychloroquine for the treatment of COVID when the authors were unable to produce the primary research data for validation due to confidentiality concerns.^5,6^

Despite legitimate barriers to sharing such as navigating intellectual property and privacy issues, and time and resource burdens^7,8^, a growing number of medical research stakeholders are calling for increased availability of data and code from cancer research. For example, funders of cancer research continue to strengthen their policies on data availability, with some like the National Institutes of Health (NIH) already mandating sharing of data under certain circumstances.^9^ Equally, a growing minority of medical journals require authors to include a data availability statement and publicly share data and code as a condition of publication.^10^ Some of these journals (e.g., Nature Cancer) even review code and software deemed integral to submitted research and use them to computationally reproduce reported findings. We also note very high levels of support from medical journal editors of requests from reviewers to access manuscripts’ raw data.^10^ Other research communities, such as the rare diseases communities, are also calling for greater availability of data to increase opportunities to re-analyse historical data to answer new research questions (i.e., secondary analyses) and combine historical data together to enhance our understanding of old ones (i.e., individual participant data meta-analyses).^11,12^ In the context of cancer research, both these types of research designs have been instrumental in shaping our understanding of topics like PSA-based screening for prostate cancer^13^ and overcoming low statistical power to reveal the benefits of treatments such as adjuvant tamoxifen for breast cancer.^14^

While significant progress has been made towards increasing the availability of the products of research (i.e., data, code and materials), previous research on the topic has reported low availability of data (0-6%) and code (0-2%) across many medical fields between 2014 and 2018.^15-25^ Furthermore, other research in medicine has shown sub-optimal compliance with journal data and code sharing policies.^26-28^ In this study we build on previous research to investigate how frequently cancer researchers share the data underlying their research, as well as the code used to perform statistical analyses in a large random sample of published cancer studies. We investigate the level of compliance with journal policies, as well as compliance with guidelines that ensure outputs are available in a way that maximally facilitates their reuse (i.e., FAIR principles^29^) – a consideration which to the authors’ knowledge has only been investigated by a single previous study in medicine.^30^ Finally, we investigate the link between some novel factors and data availability, such as the rarity of the cancer studied, the use of publicly available data in the research project and the posting of pre-prints. We also explore factors such as: the collection of data from human research subjects, open access publication models, journal impact factors and subsequent citation rates, which have been associated with data sharing and withholding in multiple fields in the past.^31,32^

## METHODS

### Study design

The following study is a cross-sectional analysis of cancer-related articles published between January 1st and December 31st, 2019. The full study protocol outlining the methods of the study was publicly registered on the Open Science Framework (OSF) on March 18th, 2020 prior to running the literature search.^33^ Important aspects of the methods are described briefly below. As the subject of study was published research articles, ethics approval was not required.

### Article selection

PubMed was searched on March 18th, 2020 to locate all oncology-related publications published in 2019. The search results were randomised in R using the sample function, then titles and abstracts, followed by full-text articles, were independently screened by two coders in parallel (DGH; JM) until the required number of eligible studies were identified. Any discrepancies between the two coders were resolved via discussion, or adjudication by another member (MJP). The eligibility criteria used for the study were as follows: 1) the article presents results of a study which recruited, involved or concerned populations, cell lines, animal analogues and/or human participants with any cancer diagnosis (benign or malignant); 2) the article was not a case report, conference abstract, synthesis of existing research (e.g. guideline, review or meta-analysis) or an opinion/news piece (e.g. editorial, letter, non-systematic expert review), 3) the article was not retracted, flagged as a duplicate publication, issued with an expression of concern, or any other reasons that would undermine trust in the research, 4) the article was written in English, available in full-text and published (electronically or in-print) between January 1st and December 31st, 2019. The full search strategy, search records, and screening results are freely available on the project’s OSF page.^33^

### Study outcomes

A comprehensive list of all the outcomes of interest to the study and their definitions are available in the study protocol and in the data dictionary on the OSF project page.^33^ The primary outcomes of interest included the public sharing of data and code. For the purposes of the study, we defined ‘code’ as the step-by-step syntax outlining all commands used within a statistical software to execute any reported analyses, and ‘data’ as the sample-level information required to reproduce and verify any or all reported qualitative or quantitative findings (including data visualisations). For example, patient-level data that would theoretically enable an independent researcher to recalculate and verify the median age of a reported cohort. In addition to the above definition of data, we also considered data sharing in the context of macromolecular structures, nucleic acid and protein sequences, microarray data and Nuclear Magnetic Resonance spectroscopy data (e.g. sharing of free induction decays^34^). However, the preparation and deposition of specimens was considered out of scope.

Two specific types of ‘sharing’ were evaluated as part of this study. The first was data and code sharing according to author declarations alone (‘reported availability’). This was defined as the presence of text, occurring anywhere in the article (e.g., in the methods section, within a formalised data/code availability statement) or supplementary material, that explicitly stated that some or all data or code underpinning the results were available, and where it can be accessed. We did not regard statements such as: ‘supplementary data are available’ or ‘data or code is available on request’ as declarations of availability. Nor did we deem references to publicly available datasets used to complement original research (e.g., to validate models) as data sharing. In the context of research that only used publicly available data (e.g., SEER data), authors needed to provide detailed information on how (or whether) the specific dataset(s) used to generate the results of the study (as opposed to the most recent iteration) could be accessed.

Secondly, data reported as available were further investigated to see determine the level of compliance with the FAIR Data guiding principles^29^ via an abbreviated version of the Australian Research Data Commons’ FAIR data self-assessment tool.^35^ Specifically, for the purposes of this study, data were considered FAIR-compliant if they were: 1) assigned both a unique and permanent identifier, 2) posted to a general, domain-specific or local institutional registry listed on re3data.org, 3) freely accessible, or accessible to researchers under explicitly stated conditions, 4) archived in a non-proprietary format (e.g., .csv, .tsv, .txt) and 5) associated with a license outlining its terms of use. Items 1-2, 3, 4 and 5 relate to the ‘Findable’, ‘Accessible’, ‘Interoperable’ and ‘Reusable’ principles respectively. If two or more datasets were posted, all were assessed and the dataset with the highest compliance was reported.

Numerous other variables were also of interest to the study, with some key variables including: 1) open access status; 2) the cancer research area classified according to the International Cancer Research Partnership’s (ICRP) ‘Common Scientific Outline’ (CSO) classification system; 3) whether the study investigated cancers classified as rare by the RARECARE project (i.e. cancers with incidence rates of less than 6 in 100,000) and 4) the number of citations accrued by each article as per Google Scholar as of April 27th, 2020 and 2021. Journal websites were also manually checked between April 28-29th, 2020 for policies governing data and code sharing, as well as the addition of availability statements.

### Data extraction

A pre-defined Google Form for data extraction was created and piloted prior to use. Primary and secondary outcome data were extracted by two authors independently in parallel (DGH; JM) for the first 198 articles (65%), with differences between coders resolved by consensus or a third party (MJP). Kappa coefficients and average percentage agreements were then calculated for each the seven primary and secondary outcome measures, following which a single author (DGH) extracted outcome data for the remaining 108 articles when inter-coder reliability was determined to be sufficiently high for the first 198 articles (kappa coefficient greater than 0.70, and the average percentage agreement greater than 95% for each domain). Refer to the OSF project page for the results of the reliability analysis.^33^

### Statistical considerations

In recognition of previous research in biomedicine that reported 13% of articles between 2015-17 both discussed and shared a functional link to research data^36^, and assuming a slightly higher estimate for the oncology literature of 15%, a random sample of 306 articles was chosen to ensure a 95% credible interval (CI) width less than 8% using the modified Jeffrey’s Interval proposed by Brown et al (2001).^37^ The method proposed is a Bayesian approach to interval estimation of binomial proportions which has been shown to provide good nominal coverage, particularly as sample proportions approach 0 (or 1).^37^

All categorical data are presented as counts and proportions. Continuous data are presented as means and standard deviations and medians and inter-quartile ranges when data were highly skewed. Credible intervals around sample proportions for binary variables using the modified Brown method were calculated using the DescTools package.^38^ In a further analysis, simple and multiple logistic regression models were also generated to estimate unadjusted and adjusted odds ratios and 95% confidence intervals for all data sharing predictors while controlling for possible confounding effects of the categorical journal data sharing policy variable (no policy, encourage, some mandatory and all mandatory). All statistical analyses were performed in R (v3.6.3).

## RESULTS

### Characteristics of included studies

The PubMed search was performed on March 18th, 2020 and yielded 200,699 records. Titles and abstracts, then full-text articles, were screened until the required 306 eligible articles were identified. Key characteristics of the 306 included studies (published in 235 unique journals) are reported in Table 1 (refer to Supplementary Table 1 for more detailed cross-tabulations).

**Table 1.**
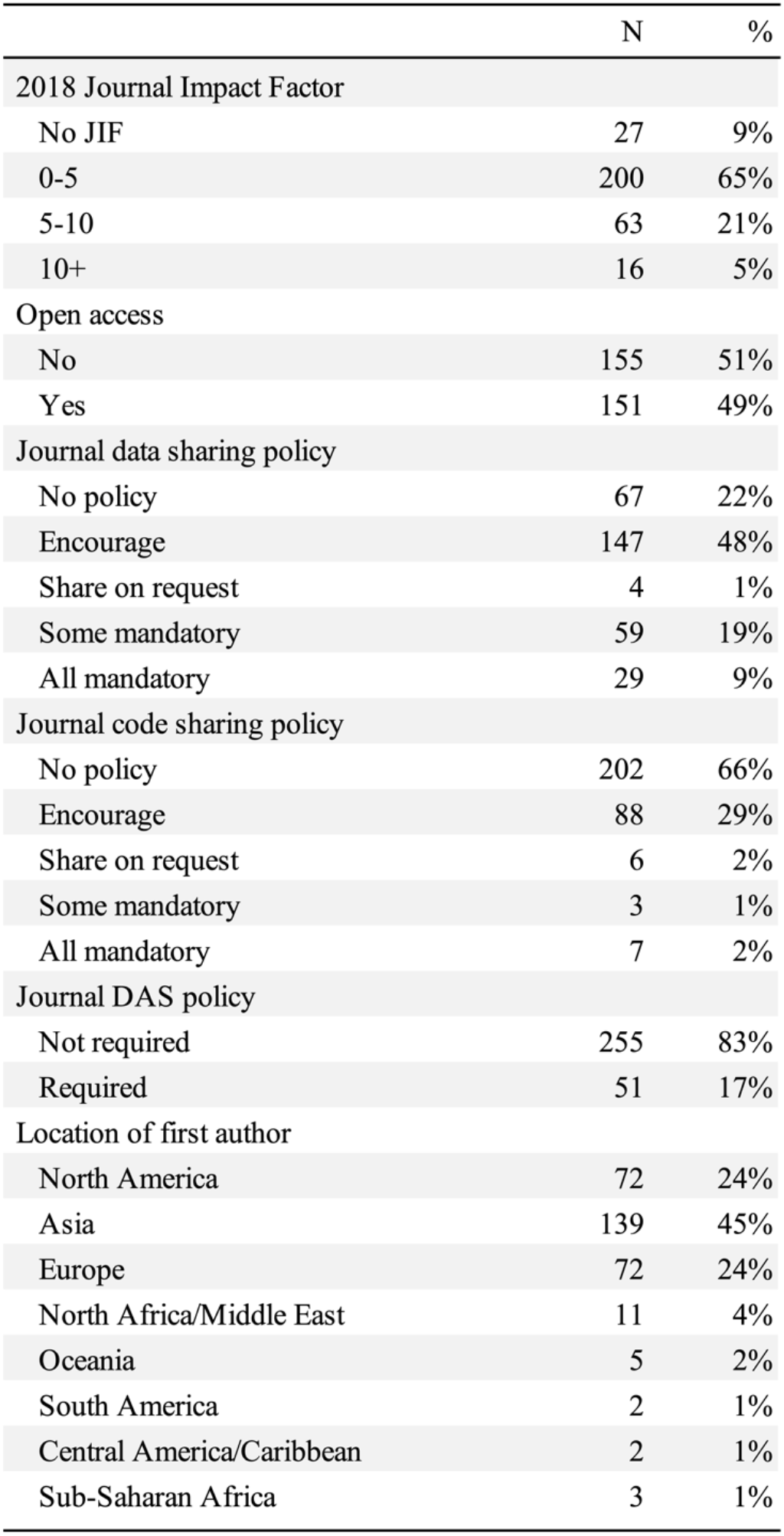
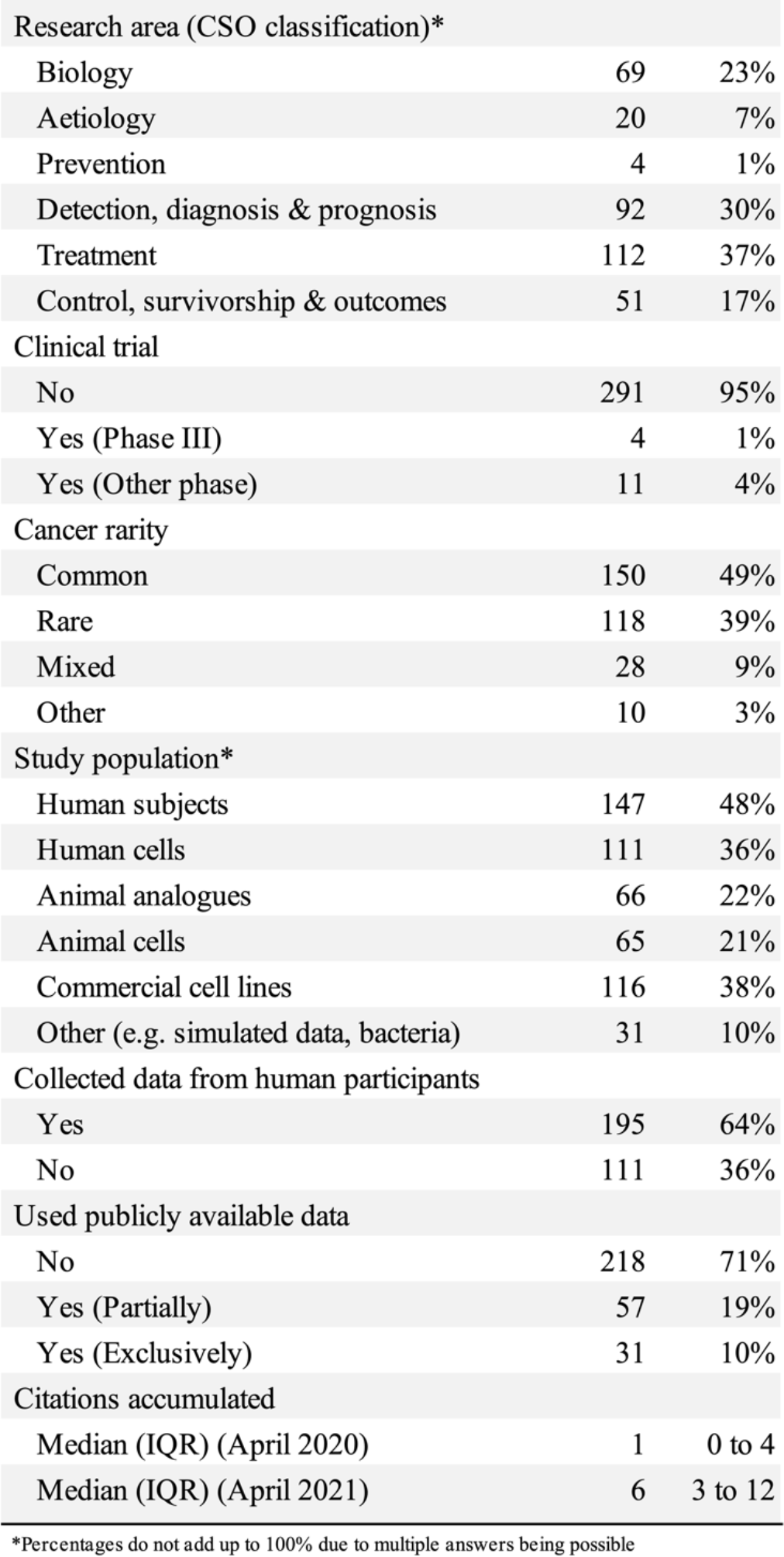
Charateristics of the included studied (N = 306)

|  | N | % |
| --- | --- | --- |
| 2018 Journal Impact Factor |  |  |
| No JIF | 27 | 9% |
| 0-5 | 200 | 65% |
| 5-10 | 63 | 21% |
| 10+ | 16 | 5% |
| Open access |  |  |
| No | 155 | 51% |
| Yes | 151 | 49% |
| Journal data sharing policy |  |  |
| No policy | 67 | 22% |
| Encourage | 147 | 48% |
| Share on request | 4 | 1% |
| Some mandatory | 59 | 19% |
| All mandatory | 29 | 9% |
| Journal code sharing policy |  |  |
| No policy | 202 | 66% |
| Encourage | 88 | 29% |
| Share on request | 6 | 2% |
| Some mandatory | 3 | 1% |
| All mandatory | 7 | 2% |
| Journal DAS policy |  |  |
| Not required | 255 | 83% |
| Required | 51 | 17% |
| Location of first author |  |  |
| North America | 72 | 24% |
| Asia | 139 | 45% |
| Europe | 72 | 24% |
| North Africa/Middle East | 11 | 4% |
| Oceania | 5 | 2% |
| South America | 2 | 1% |
| Central America/Caribbean | 2 | 1% |
| Sub-Saharan Africa | 3 | 1% |

Table 1 (cont). Characteristics of the included studies (N=306).
|  | N | % |
| --- | --- | --- |
| Research area (CSO classification)* |  |  |
| Biology | 69 | 23% |
| Aetiology | 20 | 7% |
| Prevention | 4 | 1% |
| Detection, diagnosis & prognosis | 92 | 30% |
| Treatment | 112 | 37% |
| Control, survivorship & outcomes | 51 | 17% |
| Clinical trial |  |  |
| No | 291 | 95% |
| Yes (Phase III) | 4 | 1% |
| Yes (Other phase) | 11 | 4% |
| Cancer rarity |  |  |
| Common | 150 | 49% |
| Rare | 118 | 39% |
| Mixed | 28 | 9% |
| Other | 10 | 3% |
| Study population* |  |  |
| Human subjects | 147 | 48% |
| Human cells | 111 | 36% |
| Animal analogues | 66 | 22% |
| Animal cells | 65 | 21% |
| Commercial cell lines | 116 | 38% |
| Other (e.g. simulated data, bacteria) | 31 | 10% |
| Collected data from human participants |  |  |
| Yes | 195 | 64% |
| No | 111 | 36% |
| Used publicly available data |  |  |
| No | 218 | 71% |
| Yes (Partially) | 57 | 19% |
| Yes (Exclusively) | 31 | 10% |
| Citations accumulated |  |  |
| Median (IQR) (April 2020) | 1 | 0 to 4 |
| Median (IQR) (April 2021) | 6 | 3 to 12 |

\*Percentages do not add up to 100% due to multiple answers being possible

There was a median of eight authors per article (IQR: 6-11), with 92% of first authors being affiliated with institutions in Asia (139, 45%), North America and Europe (both 72, 24%). Most articles investigated new or existing cancer treatments (112, 37%), detection, diagnostic & prognostic methods (92, 30%) or biological processes (69, 23%). Of the 306 eligible studies, 111 (36%) collected and analysed data derived only from non-human participants. The most studied cancers included breast cancer (33, 11%), bowel cancer (25, 8%), lung cancer (24, 8%), brain cancer (23, 8%), or a combination of multiple cancers (48, 16%). Almost half of the eligible articles investigated rare cancers either in isolation (118, 39%), or in combination with other common variants (28, 9%). Only 15 articles (5%) reported the results of clinical trials, four of these reporting the findings of randomised controlled trials.

### Journal characteristics

Most articles were published in subscription and gold open access journals (51% and 31% respectively), and in journals with a 2018 Impact Factor less than five (200, 65%). Almost a third of articles were subject to journal data sharing policies that either: required authors to share all data associated with the research (29, 9%), or some data under certain circumstances (59, 19%). Almost one in five articles (51, 17%) were also required by the submitting journal to complete a data availability statement. In contrast to data sharing policies, mandatory code sharing policies were much less common (10, 3%).

Other statements designed to improve transparency, such as statements outlining whether authors had any competing interests, or whether the study received funding were common (91% and 81% respectively). Similarly, most studies also declared whether ethics approval was obtained or not required (76%). In contrast, 65 (21%) and 8 (3%) articles featured formalised, stand-alone sections dedicated to addressing data and code availability respectively (i.e. data and code availability statements). Declarations of the use of open science practices, such as the public sharing of research protocols and study pre-registration were rare (2% and 4% respectively) – the latter being almost exclusively practiced by clinical trials (11/12, 92%).

### Availability of raw data

Of the 306 studies assessed, 59 declared that some or all data were publicly available (19%, 95% CI: 15-24%), 39 declared that data were available upon request (13%, 95% CI: 9-17%) and the remaining 208 articles stated that data were not available or did not provide any information on availability (68%, 95% CI: 63-73%). Of the 59 affirmatory declarations, 27 (46%) were located in dedicated data availability statements, with the remaining 32 (54%) declarations being located in other parts of the manuscript (e.g. the methods section, supplementary material). Excerpts of all 59 affirmative declarations are available on the OSF.^33^

When the 59 studies that declared data were publicly available were investigated with respect to their compliance with the FAIR principles, 49 (83%) were observed to have deposited data in a freely (or theoretically) accessible location, most commonly into data repositories (31/49, 63%) or as supplementary material on the journal website (10/49, 20%). Furthermore, when data were available for assessment, only one study (0.3%, 95% CI: 0-2%) was found to comply with the remaining four FAIR assessment criteria (Figure 1).

**Figure 1.**
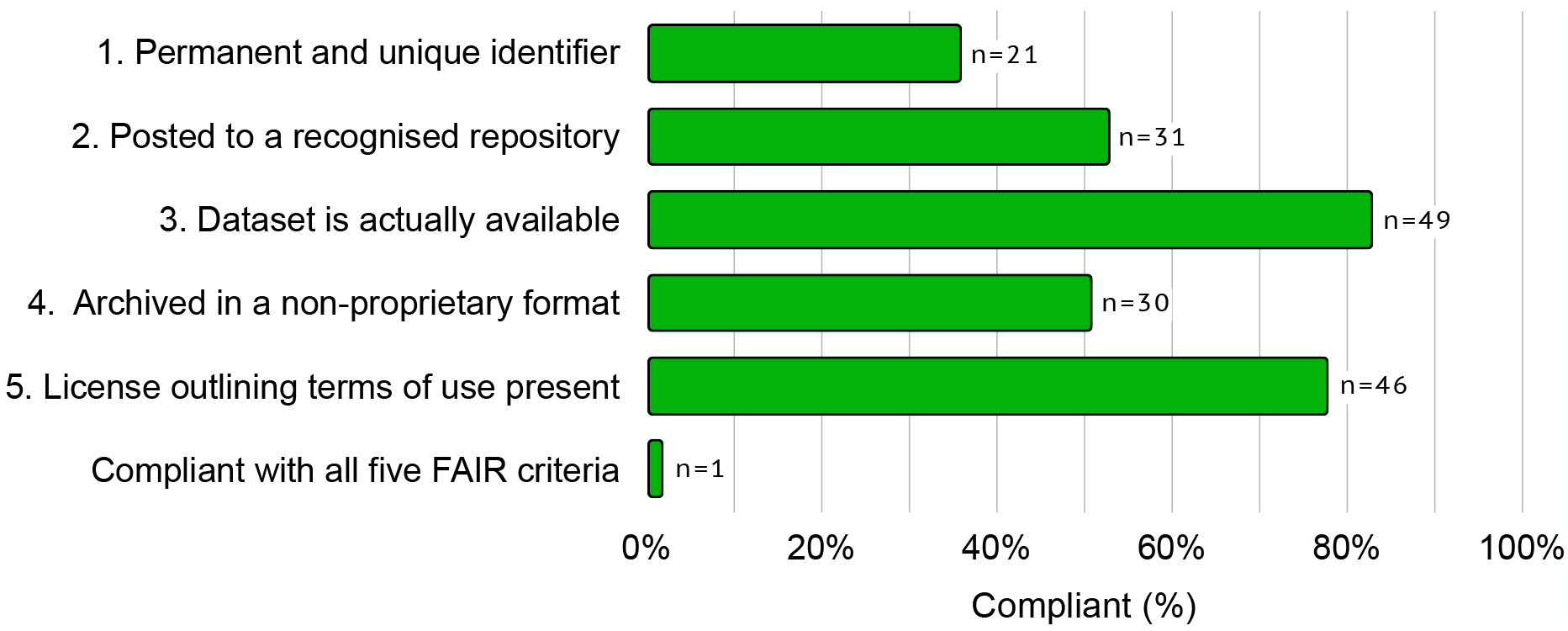
FAIR data assessment of the 59 studies that declared that data were publicly available.

The most common reasons for non-compliance included the lack of both a unique and permanent identifier (38/59, 64%), archival in a proprietary format (29/59, 49%) and not depositing data in a recognised repository (28/59, 47%).

### Statistical considerations and code sharing

Of the 306 eligible articles, 274 reported inferential statistics (90%, 95% CI: 86-93%). Of these 274 studies, ten reported that some or all code was publicly available (4%, 95% CI: 2-6%), two reported that code is available on request (1%, 95% CI: 0-2%) and 262 did not provide any information on code availability (96%, 95% CI: 93-98%). Of the ten declarations, seven originated from data or code availability statements, and the remaining three appeared in other parts of the manuscript.

Of the 274 studies that reported inferential statistics, 255 (93%, 95% CI: 90-96%) did not report performing formalised sample size calculations prior to collecting data. Furthermore, a quarter of the studies that used inferential statistics also did not report which statistical analysis software they used to analyse their data (70/274, 25%). When reported, the most frequently used software, alone or combination with others, included: SPSS (92), GraphPad (58), R (37), SAS (19) and Stata (12).

### Compliance with journal policies

Less than half of the 29 articles that were subject to a blanket mandatory data sharing policy were observed to make data available (14/29, 48%, 95% CI: 31-66%). Furthermore, of the six studies that performed inferential statistics and were subjected to a blanket mandatory code sharing policy, none reported sharing code. In contrast, 88% of articles (45/51, 95% CI: 77-95%) that were required to complete a data availability statement complied.

When comparing the effectiveness of data sharing policies, authors submitting work to journals with mandatory data sharing policies were 5.4 times more likely to share data than articles published in journals with no policy (95% CI: 2.30-12.63, p < 0.001). This association was also observed for articles published in journals that require authors to share under some circumstances but not others (RR: 2.8, 95% CI: 1.18-6.84, p = 0.013). In contrast, authors that submitted to journals that encouraged data sharing appeared to be no more likely to share data than authors publishing in journals without a data sharing policy (RR: 1.1, 95% CI: 0.43-2.65, p = 0.895). Interestingly, authors that were required to complete a data availability statement were twice as likely to share data than authors who were not (RR: 2.0, 95% CI: 1.16-3.44, p = 0.015).

### Predictors of data and code sharing

The association between sharing and withholding data with several journal and article characteristics are reported in Figure 2. Publishing in a journal with 2018 Journal Impact Factor less than ten was not associated with greater rates of sharing in comparison with journals not indexed in Clarivate’s Web of Science. In contrast, the odds of sharing data were 4.47 times (95% CI: 1.09-20.87) higher for researchers who published in a journal with an Impact Factor greater than ten compared to those who published in a journal with no impact factor. Articles in the top quartile for citations in 2021 were also associated with higher odds of data sharing rates than those in the bottom quartile (OR: 2.7, 95% CI: 1.09-7.34). However, both relationships were not found to be statistically significant when the effect of journal data sharing policies was accounted for.

**Figure 2.**
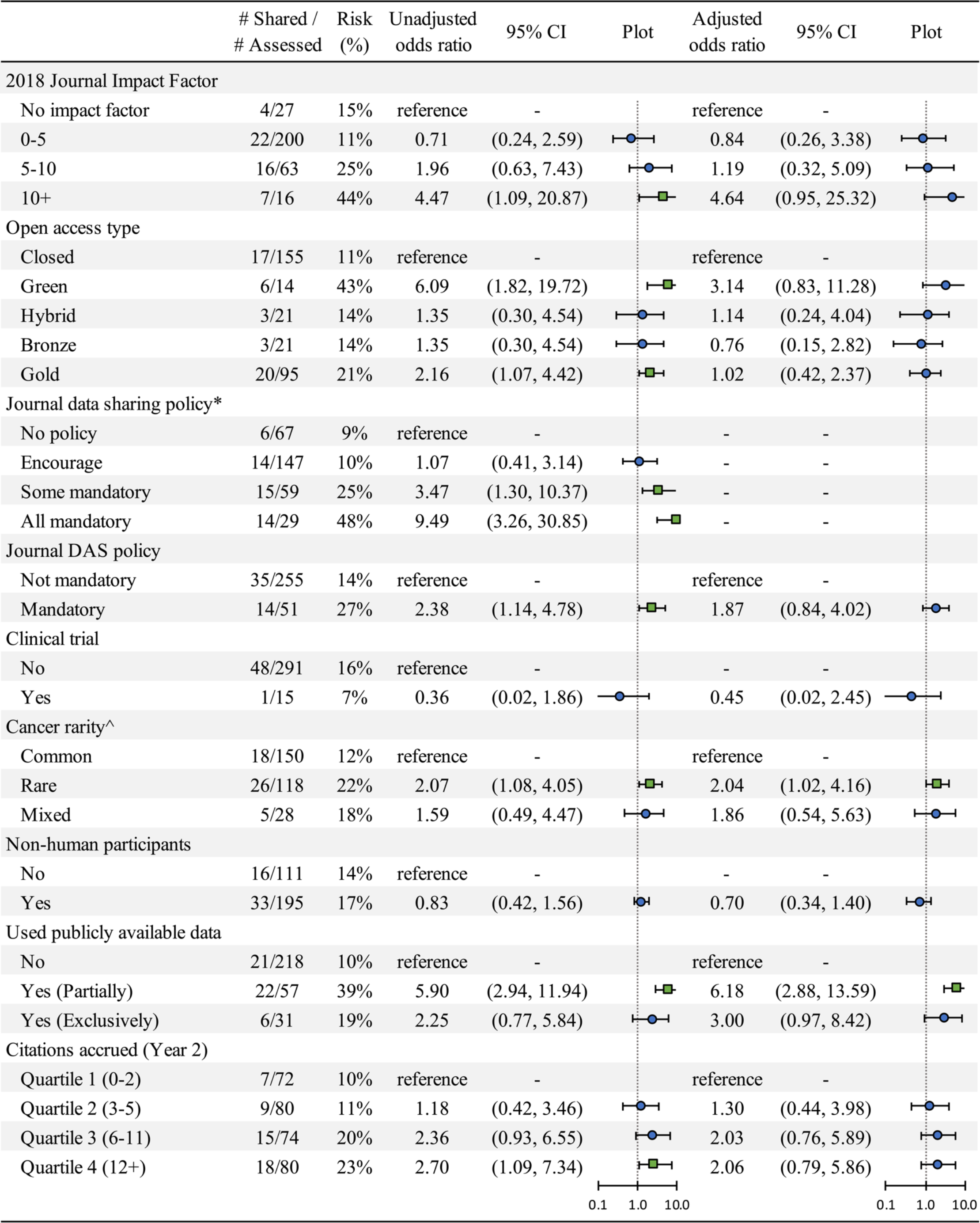
Unadjusted and adjusted odds ratios (controlling for data sharing policy) and 95% confidence intervals for the association between study characteristics and data availability. * Data from studies subject to‘share upon request’ policies (N=4) are not included. ^Data from the ‘other’ group (N=10) are not included. (Plots: Data is plotted on a algorithmic scale (Base 10), Green sqaures -*p* < 0.05; Blue circles - *p* ≥ 0.05)

The odds of sharing data were estimated to be 2.2 (95% CI: 1.07-4.42) and 6.1 times (95% CI: 1.82-19.72) higher for researchers that published in gold open access journals, or publicly shared a pre-print version of their paper both in comparison to researchers whose articles were paywalled. However, the odds of both effects halved in size when journal policies were controlled for.

The unadjusted odds of sharing data was 0.83 when researchers collected data derived from non-human participants, compared with human participants; however the precision of this estimate was relatively poor. Similarly, while we noted a 55% decrease (95% CI: 0.02-2.45) in the odds of sharing data among researchers publishing the results of clinical trials in comparison to researchers presenting the results of other study designs, even after controlling for the effects of data sharing policies, the low precision limits our ability to interpret the results conclusively. The odds that a researcher studying rare cancers or using public data to supplement their original findings shared their data were more than twice (OR: 2.1, 95% CI: 1.08-4.05) and almost six times (OR: 5.9, 95% CI: 2.94-11.94) higher respectively than those who did not. The strength and significance of both of these associations also remained stable even after controlling for the effects of data sharing policy.

Like the data sharing predictors, we noted more than a fivefold increase in the odds of reporting code to be available for researchers who shared data than for those that did not share data (OR: 5.6, 95% CI: 1.50-21.00). Additionally, the odds of reporting code to be available were 11.8 times (95% CI: 1.32-106.72) greater for researchers who publicly shared a pre-print version of their paper in comparison to researchers that published in subscription journals.

## DISCUSSION

Increasing concerns about the reliability of scientific claims continue to fuel research into the reproducibility, robustness, and generalisability of scientific findings. In modern medical research such concerns have sparked several influential research initiatives in pre-clinical medicine and cancer biology which have greatly reshaped our understanding of the extent and causes of irreproducible research^39-43^ – an issue which is of particular interest to the medical research industry given the high failure rate of clinical trials and the increasing costs of drug development and demand for more effective treatments.^40,44^

One key obstacle to reproducible research that has been highlighted by this body of research includes the overall low public availability of data, code, and materials. For example, a recent initiative by Errington and colleagues^45^ which was only able to successfully complete replications for a quarter of the shortlisted cancer biology experiments, cited low public availability of data (4/193, 2%) and code (1/78, 1%) as a major impediment (i.e., a key barrier to computing effect sizes, performing power analyses, identifying the statistical analysis strategy).

The observation of the low availability of data and code from medical research is not new. Rather our observations that 19% and 4% of cancer researchers declared data and code were publicly available respectively are consistent with several studies reporting low, but increasing, declaration rates ranging between 3-24% and 0-2% respectively over several medical disciplines between 2014-2018.^15-25^ The increase of declarations over time – particularly data availability declarations – is likely due to the growing number of medical journals that are adopting stronger policies on data and code sharing, particularly those that are requiring the addition of availability statements. For example, we note that a quarter of the unique 235 journals analysed in our study had adopted a mandatory data sharing policy for some or all data, which is higher than a previous survey of medical journal editors in the previous year.^10^ Furthermore, the proportion of articles that included a data availability statement in our study (65, 21%), which is also now a requirement for articles reporting the results of clinical trials^46^, is also consistent with prior research in medicine such as Wallach and colleagues^36^ who observed a substantial rise in the proportion of biomedical articles including an availability statement from 0% in 2009, up to 25% in 2017.

While progress is clearly being made on increasing transparency surrounding whether data is available or not, we note a large discrepancy between affirmative declarations and the sharing of data in a way that facilitates its reuse. Specifically, we noted that only one of the 59 articles that declared data was available complied with our FAIR assessment. This observation, depending on how availability for reuse is defined, is unfortunately consistent with this body of research which has reported 50-100% reductions in availability following interrogation of sharing statements^15-25^; with factors such as the lack of unique and permanent identifiers, meta-data and licensing terms being noted as major pitfalls.^30,47^ Furthermore, while we also noted a strong relationship between mandatory data sharing policies and actual data availability, we unfortunately also observed similarly sub-optimal compliance with these policies too; a finding that has been noted by other studies both inside and outside of medicine. ^26,48,49^ However, compliance issues aside, it is important to note that such policies are likely much more effective at prompting sharing than other strategies such as ‘share on request’ policies which have been associated with varying compliance rates between than 4-35%^27,50-52^, as well as encourage policies^49^ or no policy at all.^50^

In contrast to the growth of data sharing declarations over time, despite claims that code sharing is becoming increasingly normalised across many scientific fields^53^, we note persistently low code sharing rates in medicine since 2014.^15-22^ Furthermore, none of the six studies in our sample that were subject to mandatory code sharing policies reported code to be available. A finding which is consistent with the only other study to the authors’ knowledge that has examined compliance with code sharing policies in medicine by Grayling and Wheeler (2020)^28^ who reported that only 18% of the 91 methodological articles describing novel adaptive clinical trial designs that were subject to mandatory sharing policies made their code available. However, interestingly all six studies (which were also subject to mandatory data sharing policies) did address data availability, which may suggest that researchers are less aware of code sharing policies than data sharing policies. An outcome that has been documented previously in a small survey by Christian and colleagues (2020).^54^

Low compliance could also be explained by a lack of familiarity with what code sharing entails, even within the methodological research community. However, it cannot be explained by the inability to generate code given more than 90% of studies examined in both our study and that by Grayling and Wheeler^28^ used software that are all syntax-based programs (SPSS, R, SAS, Stata), or allow users to generate files that preserve the decisions made when analysing data (GraphPad).

We note some strengths of our study. First, our sample size was 4-10 times bigger than the annual estimates of previous research evaluating data and code sharing rates in medicine between 2014 and 2018.^15-22^ Random sampling of articles allowed us to make inferences about sharing rates more broadly than studies focused on sharing rates for articles published in specific journals (e.g., high impact journals), or using certain study designs (e.g., randomised controlled trials). Second, we examined entire articles and supplementary materials for declarations of data and code sharing. Third, our study is one of the very few studies to assess other factors associated with best practice archiving and sharing, such as data licensing, formatting, and discoverability, as well as assess sharing rates in the setting of rare cancers. However, we also recognise a few limitations. First, while journal policies on data and code sharing were captured shortly after running the literature search, there was still a 4 to 14-month delay between the publication of articles and the collection of this policy information. Consequently, there is a chance that some articles may have been subject to different availability policies that were superseded during this period. It is also likely that sharing rates would have further increased since 2019. This notion is supported by studies that have reported natural increases in sharing rates in other fields of medicine over time.^36^ Sharing in the modern-day context may also have been further enhanced beyond some of these estimates in the wake of global COVID-19 pandemic. Particularly as journals take on stronger positions on data availability, and as the uptake of other open science practices such as pre-printing have substantially increased.^55,56^ Both these propositions are key questions of an ongoing individual participant data meta-analysis.^57^

## CONCLUSION

Despite some progress being made towards increasing the availability of the products of research, data and code sharing in oncology has been observed to occur infrequently, and at a rate lower than would be expected if authors adhered to journal sharing policies. There is also a large gap between researchers declaring data to be available and those archiving it in a way that maximally facilitates its reuse. Both journal editors and reviewers can help with this through more active enforcement of mandatory data and code sharing policies. We also strongly encourage that researchers provide as much clarity as possible on the conditions governing access and reuse of their research data and code, even if access to such products is restricted. Additionally, we recommend that researchers and institutions consult community accepted guidelines like the FAIR principles when archiving the products of their research to maximise their value for potential reuse in the future – whether that is only by the original researchers themselves, or by other members of the cancer research community.

## Data Availability

The data, materials and code supporting the conclusions of this article are publicly available on the Open Science Framework (DOI: 10.17605/OSF.IO/Z3BFT).

https://osf.io/z3bft/

## Disclosures

### Ethics approval

Not applicable.

### Competing interests

The authors declare that they have no competing interests

### Funding

This research was supported (in part) by the Fetzer Franklin Fund of the John E. Fetzer Memorial Trust. DGH is a PhD candidate supported by an Australian Commonwealth Government Research Training Program Scholarship. MJP is supported by an Australian Research Council Discovery Early Career Researcher Award (DE200101618).

### Authors’ contributions

All authors contributed to the conception and design of the study. DGH and JM screened articles and extracted data. DGH performed all the statistical analyses and is the guarantor of the data. DGH wrote the manuscript with input from all authors. All authors read and approved the final manuscript.

## Acknowledgements

We thank Julia Milne and for their assistance with the data extraction and coding. We also thank the International Cancer Research Partnership (ICRP) for generating the CSO classifications for included publications which were provided as a collaboration between Dimensions for Funders (https://www.dimensions.ai/) and ICRP (https://www.icrpartnership.org/).

## Notes

### Competing Interest Statement

The authors have declared no competing interest.

### Clinical Protocols

https://osf.io/mjuv2

## References

1. Kong D, Ying B, Zhang J, Ying H. Retraction: The anti-osteosarcoma property of ailanthone through regulation of miR-126/VEGF-A axis. Artificial Cells, Nanomedicine, and Biotechnology 2020; 48: 1, 1254, https://doi.org/10.1080/21691401.2020.1827573.

2. Tahir M, Chaudhry EA, Zimri FK, Ahmed N, Shaikh SA, Khan S, Choudry UK, Aziz A, Jamali AR. Retraction: Negative pressure wound therapy versus conventional dressing for open fractures in lower extremity trauma. Bone Joint J 2021; 103-B(9): 1550. https://doi.org/10.1302/0301-620X.103B.BJJ-2021-00018.

3. Yuan L, Li JJ, Li CQ, et al. Retraction: Diffusion-weighted MR imaging of locally advanced breast carcinoma: the optimal time window of predicting the early response to neoadjuvant chemotherapy. Cancer Imaging 2021; 21: 62. https://doi.org/10.1186/s40644-021-00434-2.

4. Wei W, Wei X, Zhang M, Peng C. Retraction: Ultrasound microbubble-mediated miR-150-5p inhibits gastric cancer cell growth by targeting the expression of NR2F2. Journal of Receptors and Signal Transduction 2021; 41(6): 605.https://doi.org/10.1080/10799893.2021.1956794.

5. Mehra MR, Desai SS, Ruschitzka F, Patel AN. Retraction: Hydroxychloroquine or chloroquine with or without a macrolide for treatment of COVID-19: a multinational registry analysis Lancet 2020; 395. https://doi.org/10.1016/S0140-6736(20)31180-6.

6. Mehra MR, Desai SS, Kuy S, Henry TD, Patel AN. Retraction: cardiovascular disease, drug therapy, and mortality in Covid-19. N Engl J Med 2020; 382: 2582. https://doi.org/10.1056/NEJMc2021225.

7. Rathi V, Dzara K, Gross CP, Hrynaszkiewicz I, Joffe S, Krumholz HM, et al. Sharing of clinical trial data among trialists: a cross sectional survey. BMJ 2012; 345: e7570–e7570. https://doi.org/10.1136/bmj.e7570.

8. Tenopir C, Dalton ED, Allard S, Frame M, Pjesivac I, Birch B, et al. Changes in Data Sharing and Data Reuse Practices and Perceptions among Scientists Worldwide. PLOS One 2015; 10. https://doi.org/10.1371/journal.pone.0134826.

9. Contreras JL. NIH’s genomic data sharing policy: timing and tradeoffs. Trends Genet 2015; 31(2): 55–57. https://doi.org/10.1016/j.tig.2014.12.006.

10. Hamilton DG, Fraser H, Hoekstra R, et al.: Journal policies and editors’ opinions on peer review. eLife 2020; 9: e62529. https://doi.org/10.7554/eLife.62529.

11. Rubinstein YR, Robinson PN, Gahl WA, Avillach P, Baynam G, Cederroth H, Goodwin RM, Groft SC, Hansson MG, Harris NL, Huser V. The case for open science: rare diseases. JAMIA open 2020; 3(3): 472–86. https://doi.org/10.1093/jamiaopen/ooaa030.

12. Major A, Cox SM, Volchenboum SL. Using big data in pediatric oncology: Current applications and future directions. Seminars in Oncology 2020; 47: 56–64. https://doi.org/10.1053/j.seminoncol.2020.02.006.

13. Cole AP, Friedlander DF, Trinh Q. Secondary data sources for health services research in urologic oncology. Urologic Oncology: Seminars and Original Investigations; 2018. Urol Oncol 2018; 4(36). https://doi.org/10.1016/j.urolonc.2017.08.008.

14. Early Breast Cancer Trialists’ Collaborative Group. Effects of adjuvant tamoxifen and of cytotoxic therapy on mortality in early breast cancer. N Engl J Med 1988; 319(26): 1681–92. https://doi.org/10.1056/NEJM198812293192601.

15. Anderson JM, Wright B, Rauh S, Tritz D, Horn J, Parker I, et al. Evaluation of indicators supporting reproducibility and transparency within cardiology literature. Heart 2021; 107: 120–6. https://doi.org/10.1136/heartjnl-2020-316519.

16. Smith CA, Nolan J, Tritz DJ, Heavener TE, Pelton J, Cook K, et al. Evaluation of reproducible and transparent research practices in pulmonology. Pulmonology 2021; 27: 134–43. https://doi.org/10.1016/j.pulmoe.2020.07.001.

17. Fladie IA, Evans S, Checketts J, Tritz D, Norris B, Vassar BM. Can Orthopaedics become the Gold Standard for Reproducibility? A Roadmap to Success. bioRxiv 2019. https://doi.org/10.1101/715144.

18. Rauh S, Torgerson T, Johnson AL, Pollard J, Tritz D, Vassar M. Reproducible and transparent research practices in published neurology research. Research Integrity and Peer Review 2020; 5: 5. https://doi.org/10.1186/s41073-020-0091-5.

19. Wright BD, Vo N, Nolan J, Johnson AL, Braaten T, Tritz D, et al. An analysis of key indicators of reproducibility in radiology. Insights into Imaging 2020; 11: 65. https://doi.org/10.1186/s13244-020-00870-x.

20. Sherry CE, Pollard JZ, Tritz D, Carr BK, Pierce A, Vassar M. Assessment of transparent and reproducible research practices in the psychiatry literature. General Psychiatry 2020; 33: e100149. https://doi.org/10.1136/gpsych-2019-100149.

21. Evans S, Fladie I, Anderson M, Tritz D, Vassar M. Evaluation of Reproducible and Transparent Research Practices in Sports Medicine Research: A Cross-sectional study. bioRxiv 2019. https://doi.org/10.1101/773473.

22. Walters C, Harter ZJ, Wayant C, Vo N, Warren M, Chronister J, et al. Do oncology researchers adhere to reproducible and transparent principles? A cross-sectional survey of published oncology literature. BMJ Open 2019; 9: e033962. https://doi.org/10.1136/bmjopen-2019-033962.

23. Rauh SL, Johnson BS, Bowers A, Tritz D, Vassar M. Evaluation of Reproducibility in Urology Publications. bioRxiv 2019. https://doi.org/10.1101/773945.

24. Adewumi MT, Vo N, Tritz D, Beaman J, Vassar M. An evaluation of the practice of transparency and reproducibility in addiction medicine literature. Addictive Behaviors 2021; 112: 106560. https://doi.org/10.1016/j.addbeh.2020.106560.

25. Fladie IA, Adewumi TM, Vo NH, Tritz DJ, Vassar MB. An Evaluation of Nephrology Literature for Transparency and Reproducibility Indicators: Cross-Sectional Review. Kidney International Reports 2020; 5: 173–81. https://doi.org/10.1016/j.ekir.2019.11.001.

26. Alsheikh-Ali AA, Qureshi W, Al-Mallah MH, Ioannidis JPA. Public Availability of Published Research Data in High-Impact Journals. PLOS ONE 2011; 6: e24357. https://doi.org/10.1371/journal.pone.0024357.

27. Rowhani-Farid A, Barnett AG. Has open data arrived at the British Medical Journal (BMJ)? An observational study. BMJ Open 2016; 6: e011784. https://doi.org/10.1136/bmjopen-2016-011784.

28. Grayling MJ, Wheeler GM. A review of available software for adaptive clinical trial design. Clin Trials 2020; 17: 323–31. https://doi.org/10.1177/1740774520906398.

29. Wilkinson M, Dumontier M, Aalbersberg I, et al. The FAIR Guiding Principles for scientific data management and stewardship. Sci Data 2016; 3, 160018. https://doi.org/10.1038/sdata.2016.18.

30. Zuo X, Chen Y, Ohno-Machado L, Xu H. How do we share data in COVID-19 research? A systematic review of COVID-19 datasets in PubMed Central Articles. Brief Bioinform 2021; 22: 800–11. https://doi.org/10.1093/bib/bbaa331.

31. Piwowar HA (2011) Who Shares? Who Doesn’t? Factors Associated with Openly Archiving Raw Research Data. PLOS One 6(7): e18657. https://doi.org/10.1371/journal.pone.0018657.

32. Piwowar HA, Vision TJ. Data reuse and the open data citation advantage. PeerJ 2013; 1: e175. https://doi.org/10.7717/peerj.175.

33. Hamilton DG, Fidler F, Page MJ. How common is data and code sharing in the oncology literature? Open Science Framework 2020. https://doi.org/10.17605/OSF.IO/Z3BFT.

34. Bisson J, Simmler C, Chen S-N, Friesen JB, Lankin DC, McAlpine JB, et al. Dissemination of original NMR data enhances reproducibility and integrity in chemical research. Nat Prod Rep 2016; 33: 1028–33. https://doi.org/10.1039/C6NP00022C.

35. Levett K, Russell K, Schweitzer M, Unsworth K, White A. (2021), FAIR Data Assessment Tool (v1.0), GitHub repository, https://github.com/au-research/FAIR-Data-Assessment-Tool. [Last accessed: 18 Nov 2021.]

36. Wallach JD, Boyack KW, Ioannidis JPA. Reproducible research practices, transparency, and open access data in the biomedical literature, 2015–2017. PLOS Biol 2018; 16: e2006930. https://doi.org/10.1371/journal.pbio.2006930.

37. Brown LD et al. Interval Estimation for a Binomial Proportion. Statistical Science 2001; 16(2): 101–17.

38. Signorell A. DescTools: Tools for descriptive statistics. 2019. R package version 0.99.29. https://CRAN.R-project.org/package=DescTools.

39. Prinz F, Schlange T, Asadullah K. Believe it or not: how much can we rely on published data on potential drug targets? Nat Rev Drug Discov 2011; 10(9): 712. https://doi.org/10.1038/nrd3439-c1.

40. Begley CG, Ellis LM. Drug development: Raise standards for preclinical cancer research. Nature 2012; 483(7391): 531–3. https://doi.org/10.1038/483531a.

41. Perrin S. Preclinical research: Make mouse studies work. Nature 2014; 507(7493): 423–425. https://doi.org/10.1038/507423a.

42. Amaral OB, Neves K, Wasilewska-Sampaio AP, Carneiro CF. The Brazilian Reproducibility Initiative. eLife 2019; 8: e41602. https://doi.org/10.7554/eLife.41602.

43. Errington TM, Mathur M, Soderberg CK, Denis A, Perfito N, Iorns E, Nosek BA. Investigating the replicability of preclinical cancer biology. eLife 2021; 10: e71601. https://doi.org/10.7554/eLife.71601.

44. Wong CH, Siah KW, Lo AW. Corrigendum: Estimation of clinical trial success rates and related parameters, Biostatistics 2019; 20(2): 366. https://doi.org/10.1093/biostatistics/kxy072.

45. Errington TM, Denis A, Perfito N, Iorns E, Nosek BA. Challenges for assessing replicability in preclinical cancer biology. eLife 2021; 10: e67995. https://doi.org/10.7554/eLife.67995.

46. Taichman DB, Sahni P, Pinborg A, Peiperl L, Laine C, James A, et al. Data Sharing Statements for Clinical Trials - A Requirement of the International Committee of Medical Journal Editors. N Engl J Med 2017; 376: 2277–9. https://doi.org/10.1056/NEJMe1705439.

47. Tuyl SV, Whitmire AL. Water, Water, Everywhere: Defining and Assessing Data Sharing in Academia. PLOS One 2016; 11: e0147942. https://doi.org/10.1371/journal.pone.0147942.

48. Vines TH, Andrew RL, Bock DG, Franklin MT, Gilbert KJ, Kane NC, et al. Mandated data archiving greatly improves access to research data. FASEB J 2013; 27: 1304–8. https://doi.org/10.1096/fj.12-218164.

49. Piwowar, H., Chapman, W. A review of journal policies for sharing research data. Nat Prec 2008. https://doi.org/10.1038/npre.2008.1700.1.

50. Gabelica M, Cavar J, Puljak L. Authors of trials from high-ranking anesthesiology journals were not willing to share raw data. J Clin Epidemiol 2019; 109: 111–6. https://doi.org/10.1016/j.jclinepi.2019.01.012.

51. Wicherts JM, Borsboom D, Kats J, Molenaar D. The poor availability of psychological research data for reanalysis. Am Psychol 2006; 61(7): 726–8. https://doi.org/10.1037/0003-066X.61.7.726.

52. Savage CJ, Vickers AJ. Empirical Study of Data Sharing by Authors Publishing in PLOS Journals. PLOS One 2009; 4: e7078. https://doi.org/10.1371/journal.pone.0007078.

53. Goldacre B, Morton CE, DeVito NJ. Why researchers should share their analytic code. BMJ 2019: l6365. https://doi.org/10.1136/bmj.l6365.

54. Christian T-M, Gooch A, Vision T, Hull E. Journal data policies: Exploring how the understanding of editors and authors corresponds to the policies themselves. PLOS One 2020; 15(3): e0230281. https://doi.org/10.1371/journal.pone.0230281.

55. Abdill RJ, Blekhman R. Tracking the popularity and outcomes of all bioRxiv preprints. eLife 2019; 8: e45133. https://doi.org/10.7554/eLife.45133.

56. Fraser N, Brierley L, Dey G, Polka JK, Pálfy M, Nanni F, et al. The evolving role of preprints in the dissemination of COVID-19 research and their impact on the science communication landscape. PLOS Biology 2021; 19: e3000959. https://doi.org/10.1371/journal.pbio.3000959.

57. Hamilton DG, Fraser H, Fidler F, et al. Rates and predictors of data and code sharing in the medical and health sciences: Protocol for a systematic review and individual participant data meta-analysis. [version 2; peer review: 2 approved]. F1000Res 2021; 10: 491. https://doi.org/10.12688/f1000research.53874.2.

